# IDH and 1p19q Diagnosis in Diffuse Glioma from Preoperative MRI Using Artificial Intelligence

**DOI:** 10.1101/2023.04.26.21267661

**Authors:** Hugh McHugh, Soroush Safaei, Gonzalo D. Maso Talou, Stephen L. Gock, Joo Yeun Kim, Alan Wang

## Abstract

**Background:** Isocitrate dehydrogenase (IDH) mutation and 1p19q codeletion are important beneficial prognosticators in glioma. IDH and 1p19q diagnosis requires tissue sampling and there are likely benefits of presurgical diagnosis. Research supports the potential of MRI-based IDH and 1p19q diagnosis, however there is a paucity of external validation outside the widely used The Cancer Imaging Archive (TCIA) dataset. We present a combined IDH and 1p19q classification algorithm and assess performance on a local retrospective cohort (NZ) and the Erasmus Glioma Database (EGD).

**Methods:** 2D convolutional neural networks are trained to provide IDH and 1p19q classification. Inputs are T1 post-contrast, T2, and FLAIR sequences. Training data consists of preoperative imaging from the TCIA dataset (n=184) and a locally obtained NZ dataset (n=349). Evaluation data consists of the most recent cases from the NZ dataset (n=205) and the EGD (n=420).

**Results:** IDH classification accuracy was 93.3% and 91.5% on the NZ and EDG, with AUC values of 95.4% and 95.8%, respectively. 1p19q accuracy was 94.5% and 87.5% with AUC values of 92.5% and 85.4% on the NZ and EGD datasets. Combined IDH and 1p19q accuracy was 90.4% and 84.3% on the NZ and EGD, with AUC values of 92.4% and 91.2%.

**Conclusions:** High IDH and 1p19q classification performance was achieved on the NZ retrospective cohort. Performance generalised to the EGD demonstrating the potential for clinical translation. This method makes use of readily available imaging and has high potential impact in glioma diagnostics.

**Key Points:**

- IDH and 1p19q are the main molecular markers in glioma.
- Accurate predictions can be obtained from preoperative MRI without changes to imaging protocols.
- Non-invasive diagnosis will likely enhance treatment planning and facilitate targeted preoperative therapies.

**Importance of the Study:** The 2021 WHO CNS tumour classification system formalises the increasing recognition of molecular factors like IDH and 1p19q in the prognostication and treatment of glioma. Emerging research shows the potential of artificial intelligence methods applied to preoperative MRI sequences to noninvasively predict molecular status. A limitation of the literature published to date is a lack of generalisation and external validation outside the widely used TCIA dataset. Here we present the performance of an MRI-based IDH and 1p19q classification tool evaluated on a large consecutive cohort from New Zealand and an independent publicly available dataset of MR images from the Netherlands. We demonstrate high predictive performance with robust generalisation, indicating the potential usefulness of this method in the workup of glioma. Reliable preoperative tumour characterisation may facilitate tailored treatment approaches and early decision making without the need for additional imaging.

## Introduction

Glioma is the most common primary brain malignancy with 33,000 new diagnoses of glioma in the USA annually [1]. Unlike circumscribed gliomas, tumour cells in diffuse glioma infiltrate into the surrounding tissues, which means they generally recur following resection. Treatment of diffuse glioma consists of surgery followed by a combination of chemotherapy and radiotherapy. Although patients with genetically favorable tumours may survive for many years, these tumours inevitably undergo malignant transformation, and patients are at risk of long-term disease and treatment-related complications.

IDH is either mutated or wild-type and IDH mutation confers a considerable survival benefit. 1p19q is either codeleted or intact, where codeletion refers to the chromosomal deletion of the p-arm of chromosome 1 and the q-arm of chromosome 19. 1p19q codeletion exists in a subset of IDH mutated tumours and these patients have additional survival benefits over IDH mutation alone. Reported survival varies, but median survival has been reported at 20 months and 65 months for IDH wild-type vs mutant tumours respectively [2]. Patients with IDH mutated - 1p19q codeleted tumours may outlive study follow-up periods and 5-year survival rates of 74% have been reported [3].

Following the recognition of IDH mutation in glioma in 2008 [4], there has been increasing emphasis on IDH mutation and 1p19q codeletion over histological grade as key determinants of clinical outcomes. This is reflected in revisions to the World Health Organisation (WHO) CNS tumour classification system with the introduction of glioma subcategories for IDH status in 2016. At this point, 1p19q codeletion became an entity defining feature of oligodendroglioma. These changes were driven by the strong observed effect of tumour genetics on patient outcomes [5]. The 2021 WHO classification places further emphasis on IDH status. Lack of IDH mutation is now the key differentiating factor for glioblastoma (GBM), so grade 4 IDH mutant tumours are not GBMs and all IDH wild-type diffuse gliomas are GBMs regardless of grade [6].

IDH mutation results in a distinct tumour pathophysiology which is related to the production of D-2-hydroxyglutarate in cancer cells [7]. IDH mutated tumours are more commonly low-grade, with a prevalence of IDH mutation of approximately 60% in grade 2 and 3 tumours compared to 10% in grade 4 tumours [8]. The overall prevalence of IDH mutation varies across clinical datasets but is close to 30% [9]. This correlates with the higher incidence of high-grade gliomas over low-grade gliomas [10,11]. Roughly 50% of IDH mutated tumours are 1p19q codeleted. 1p19q codeletion does not occur in IDH wild-type tumours.

At present, IDH diagnosis is performed by immunohistochemistry or genetic sequencing of tissue samples, 1p19q codeletion detection is made by fluorescence in situ hybridisation (FISH). Both approaches require either a surgical brain biopsy or tumour resection. Several methods of noninvasive IDH diagnosis have been proposed, however none are in routine practice. These include MR spectroscopy, biochemical detection of 2-hydroxyglutarate and isolating tumour DNA from CSF [12,13,14]. 1p19q codeletion does not result in specifically recognised biochemical changes making non-invasive detection more difficult. Radiologically, the FLAIR-T2 mismatch sign is highly specific for IDH mutation with specificity of 98% but low sensitivity of 26% [15]. Microcystic change within the tumour has been proposed as the mechanism [16]. 1p19q codeleted oligodendrogliomas commonly demonstrate calcification on imaging [17].

A promising method is the application of Artificial Intelligence (AI) methods on MRI data. MRI is the modality of choice for the workup of brain tumours. Tumour protocols universally include T2, FLAIR, and T1 pre- and post-contrast sequences [18]. An advantage of this method is that it utilises already available data and does not require additional scanning time or modifications to MRI protocols.

A growing body of research is validating MRI based molecular diagnosis, and high performance has been reported using convolutional neural networks on processed MRI using standard sequences only [19-26]. Much of the research to date has utilised publicly available MRI data, principally from TCIA [27]. External validation of these methods has been performed (Choi and others), however, high performance for IDH prediction has generally not been replicated on external datasets. Generalisability is a central concern in the clinical translation of this technology as performance tends to fall when AI methods are applied to new data. This is particularly true of MRI where technical factors have large effects on quantitative aspects of the final image. Verifying the robustness of molecular prediction models on external datasets is a crucial step in the clinical translation of this technology.

There are several anticipated benefits of knowing IDH and 1p19q status before surgical resection. Preoperative molecular diagnosis may assist neurosurgeons decide on the optimal surgical approach. Evidence suggests that patients with IDH mutated tumours benefit from a more aggressive resection which includes non-enhancing tissue [28]. There are also several IDH specific chemotherapeutic agents in development. Given that reduction in tumour volume has been demonstrated with chemotherapy, neoadjuvant chemotherapy is a potential tool to improve tumour resectability [29]. Accurate preoperative molecular diagnosis might allow the investigation of targeted neoadjuvant therapies. Accelerated prognostication would also better inform decisions about treatment and end-of-life planning for patients with this devastating disease.

Here we present a combined IDH and 1p19q prediction model evaluated on a local retrospective clinical cohort, as well as a large external cohort. Because IDH and 1p19q status are the key differentiating factors in the 2021 WHO CNS classification system [6], our method can classify tumours as GBM, IDH mutated astrocytoma, and IDH mutated - 1p19q codeleted oligodendroglioma.

## Materials and Methods

Ethics approval was obtained via the Auckland Health Regional Ethics Committee (AH3229) as well as locality approval from Auckland District Health Board (A+9022) prior to commencement.

### Participants

The New Zealand (NZ) dataset consists of a consecutive cohort of patients presented at the Auckland City Hospital Neuropathology Multidisciplinary Meeting between 2011 and 2021. This represents all adult tumour cases operated on over this period by the Department of Neurosurgery for whom histopathology was performed. There were a total of 752 patients with glioma presented over this period. Inclusion criteria for the final imaging analysis were: availability of preoperative MRI, histologically proven diffuse glioma (WHO grade 2-4) with IDH status and 1p19q status if IDH mutant, and age at least 18 years. Patients without suitable imaging, confirmed glioma with mutation status or younger than 18 were excluded. Following the application of inclusion criteria, 554 cases were included in the NZ dataset. The NZ dataset was split according to the date of the preoperative MRI. Scans obtained prior to 01/01/2018 were included in the NZ training dataset (*n* = 349) and those obtained between 01/01/2018 and 31/08/2021 were included in the NZ testing dataset (*n* = 205). The most recent cases were chosen as the NZ testing data as this allows a simulated clinical scenario, whereby performance would be as expected had the model been trained prior to this period and evaluated prospectively.

Training data was supplemented by 184 MR images from TCIA dataset [30,31,27]. These cases were screened by the same criteria.

The external testing data consisted of the Erasmus Glioma Database (EGD) screened by the above criteria (*n* = 420) [32]. All cases with available IDH status, 1p19q codeletion status if IDH mutant, and age were included in this analysis. Ground truth in all cases is histological IDH and 1p19q status. A list of the EDG subjects used has been provided in the Supplementary Material for reproducibility.

### Experimental Design

2D Dense U-Nets were trained on the training data and evaluated on the NZ and EGD testing cohorts. Training and evaluation were repeated three times to account for model stochasticity.

### Image Preprocessing

Images in the NZ and TCIA datasets were processed to ensure data consistency. MRI sequences were converted using dcm2niix v1.0 [33]. Rigid registration was performed using Simple ITK v2.0 [34,35,36] to the SRI-24 Atlas [37]. Sequences were resampled to a uniform 1mm^3^ isotropic resolution with an image size of 240×240×155 voxels. Images were normalised using z-scoring, i.e.,

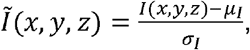

where *μ*_*l*_ and *σ*_*l*_ are the mean and standard deviation value of original image *I*. Brain extraction and tumour segmentation was performed using separate 2D Dense U-Nets trained on the BraTS 2021 training data [38,39,40].

MRIs from the EGD are obtained preprocessed as detailed in [32]. Preprocessing is similar with 1 mm^3^ resolution, the principle difference is that the EGD is registered to the MNI152 atlas [41,42]. We perform z-scoring normalisation as described above as an additional preprocessing step.

### Model Implementation

A 2D Dense U-Net was implemented in Tensorflow v2.0 [43]. This is a variation of the U-Net with the addition of dense-blocks [44,45]. The U-Net architecture consists of an encoder and a decoder with skip connections (see Figure 1). The Dense modification involves concatenating the feature map of the output of each layer with the initial input [45].

**Figure 1:**
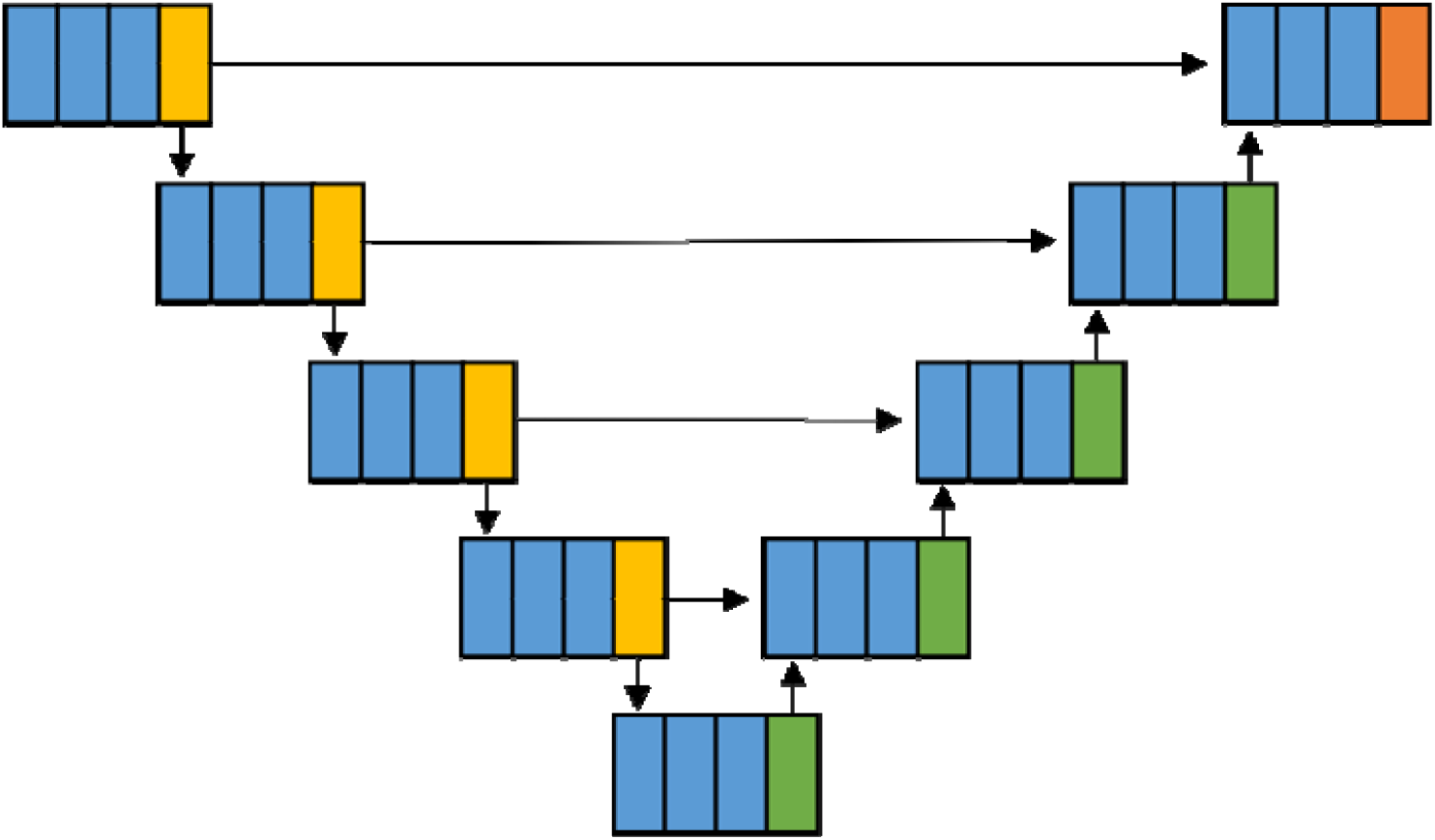
Diagram of the model architecture. Dense-blocks are blue, transition-down blocks are gold, transition-up blocks are green, the activation layer is orange, and skip connections are represented by arrows.

The encoder consists of four sequential layers of increasing depth which are composed of three dense-blocks in series followed by a transition-down block which utilises max pooling to downsample the feature map. The bottleneck consists of three dense-blocks in series. The decoder has four layers of reducing depth which is made up o a transition-up block followed by three dense-blocks. The output of each encoder layer is concatenated with the input of each corresponding decoder layer. The final layer is a convolution which provides the voxel-wise Softmax activation of each segmentation label.

### Model Inputs and Outputs

The IDH and 1p19q classification models were trained on axial slices consisting of the T2, FLAIR, and T1ce sequences with an image size of. Age is added as an additional input feature which is concatenated as a constant with an image size corresponding to the feature map at each level of the decoder. Models were trained with tumour segmentation masks as targets to predict image voxels in 4 mutually exclusive classes: (i) no tumour, (ii) IDH wild-type, (iii) IDH mutated - 1p19q intact or (iv) IDH mutated - 1p19q codeleted.

Segmentation volumes corresponding to the input MRI are obtained by iterating over the slices of the MRI volume. Two-stage classification of the tumour is performed. IDH classification is obtained using a majority vote approach based on the number of voxels in the image classified as either IDH wild-type or mutated (1p19q intact or codeleted). If the tumour is predicted as IDH mutated, 1p19q classification is made via majority vote of IDH mutated voxels predicted as 1p19q intact versus codeleted.

The IDH and 1p19q segmentations can be represented as colour images where colour channels correspond to voxels predicted as IDH wild-type, IDH mutated - 1p19q intact, and IDH mutant - 1p19q codeleted. Figure 2 demonstrates an example segmentation of each tumour subtype from the NZ testing dataset. The IDH confidence score is the proportion of voxels predicted as IDH mutated, this serves as a certainty metric for model prediction.

**Figure 2:**
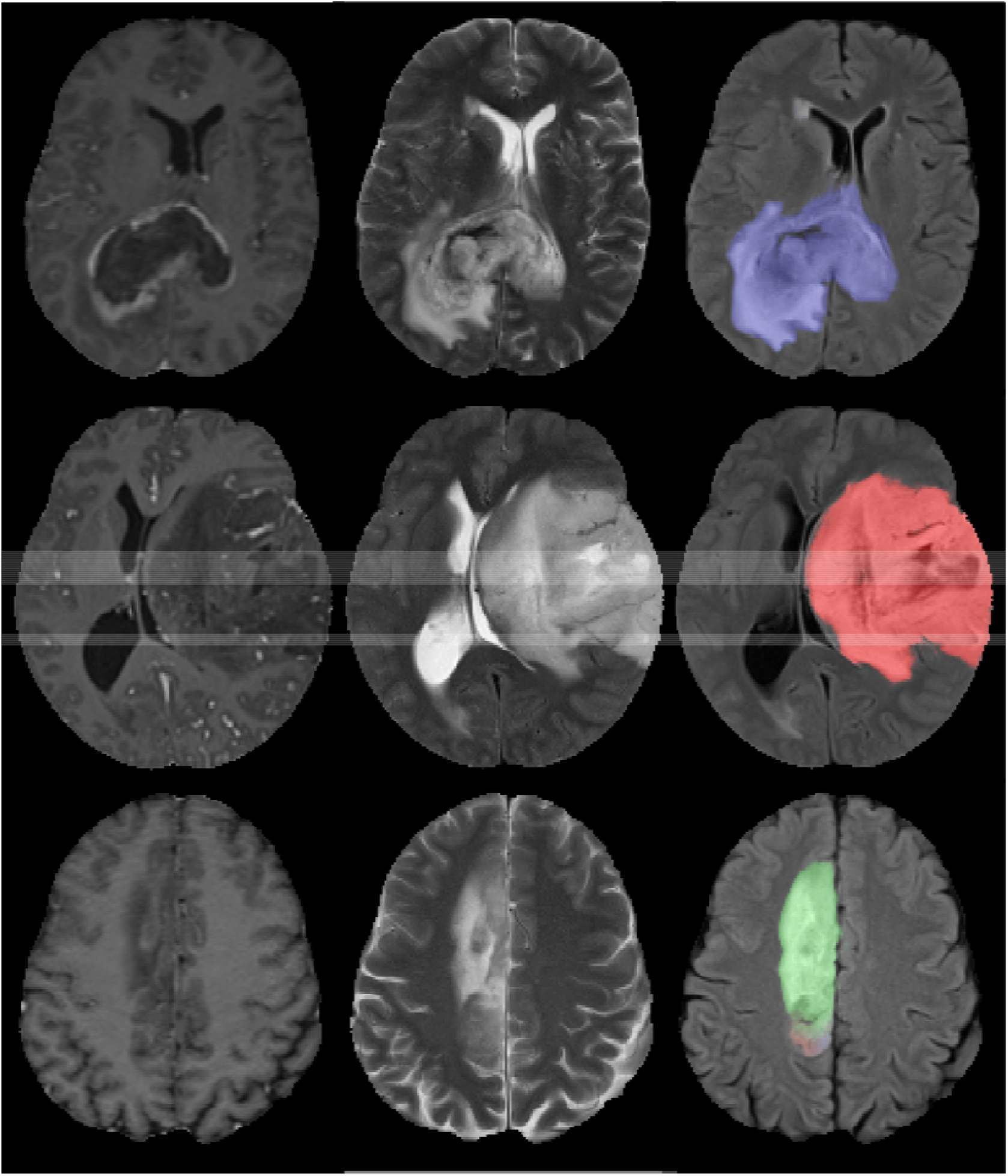
Images and IDH/1p19q segmentations from the NZ testing dataset. From left to right: T1 post-contrast, T2 and FLAIR with segmentation overlay. From top to bottom: IDH wild-type GBM (blue), IDH mutant - 1p19q intact astrocytoma (red), IDH mutant - 1-19q codeleted oligodendroglioma (green).

### Training

Augmentation consisted of shifting the intensity of the whole slice by a uniform amount, addition of Gaussian noise, random translation, random rotation, and left-right flipping. The loss function was weighted cross-entropy, and the model was trained using the Adam optimiser with batch size of 8, and a learning rate of. Model training was terminated after 200 epochs. Model weights from epoch 200 were used for model evaluation.

Model training occurred on a 32GB NVIDIA Tesla V100 GPU. The training time was 165 h per experiment. Prediction times for each subject were less than 3 s on the GPU. Generation of IDH prediction segmentation requires less than 6 min of processing on a standard non-GPU laptop.

### Statistical Analysis

All results reported represent subject-wise classification and are presented as the mean over the three repeated experiments plus or minus one standard deviation from the mean. Proportions were compared with the Chi-squared test and means were compared using analysis of variance (ANOVA). Statistical analyses were performed using SPSS. Area under the receiver operator curve (AUC) was calculated using the Scikit-Learn Python package [46].

## Results

### Demographics and Molecular Status

There were significant differences in age between the datasets with no difference in sex. Patients in the TCIA dataset were younger than the NZ and EGD datasets. There were significant differences in proportions of each molecular subtype with increased rates of IDH mutation in the TCIA and EGD datasets compared to the NZ training and testing datasets. Table 1 displays the demographic and molecular characteristics of the training and testing datasets.

**Table 1:** Patient demographics and molecular characteristics of each dataset.

|  | Training |  | Testing |  | p-value |
| --- | --- | --- | --- | --- | --- |
|  | TCIA (n = 184) | NZ (n = 349) | NZ (n = 205) | EGD (n = 420) |  |
| **Mean Age** | 52.3 ± 15.1 | 56.2 ± 14.8 | 56.6 ± 14.7 | 55.9 ± 15.1 | 0.015 |
| **Sex** |  |  |  |  | 0.076 |
| Female | 90 (48.9%) | 135 (38.7%) | 88 (42.9%) | 162 (38.6%) |  |
| Male | 94 (51.1%) | 214 (61.3%) | 117 (57.1%) | 258 (61.4%) |  |
| **Molecular Status** |  |  |  |  | < 0.001 |
| IDH Wild-Type | 106 (57.6%) | 278 (79.7%) | 173 (84.4%) | 306 (72.9%) |  |
| IDH Mutated | 78 (42.4%) | 71 (20.3%) | 32 (15.6%) | 114 (27.1%) |  |
| - 1p19q Intact | 53 (28.8%) | 31 (8.8%) | 18 (8.8%) | 43 (10.2%) |  |
| - 1p19q Codeleted | 25 (13.6%) | 40 (11.5%) | 14 (6.8%) | 71 (16.9%) |  |

### Image Characteristics

There were significant differences in mean slice thickness across the datasets for all sequences. Here, volumetric imaging was considered as sequences acquired with a slice thickness of less than 1.5 mm. Slice thickness for T1 post-contrast was thinner in the NZ training and NZ testing datasets with increasing utilisation of volumetric T1 post-contrast sequences from the training to the testing datasets. Volumetric T2 sequences were not present in any of the datasets. Increasing frequency of volumetric FLAIR sequences were seen in the NZ training and NZ testing datasets, respectively. Table 2 demonstrates the imaging characteristics of the TCIA and NZ datasets. Slice thickness information was not available for the EGD dataset.

**Table 2:**
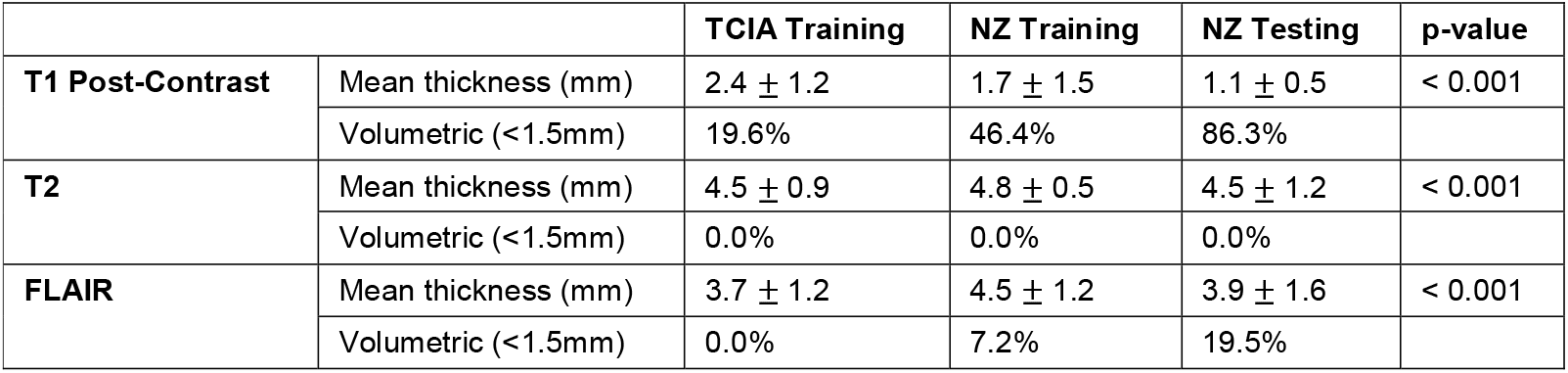
Slice thickness and proportion of volumetrically acquired sequences for each sequence in TCIA and NZ datasets.

### Model Performance

IDH prediction accuracy on the NZ testing data was 93.3% with 69.8% sensitivity and 97.7% specificity. AUC was 95.4%. On the external EGD dataset, accuracy was 91.5% with 82.6% sensitivity and 96.1% specificity. AUC was 95.8%.

1p19q predictive accuracy on the NZ testing data was 94.5% with 59.5% sensitivity and 97.0% specificity. AUC was 92.5%. Corresponding 1p19q performance values on the EGD dataset were: 87.5%, 54.0%, 94.4% and 85.4%. The input score for the 1p19q AUC calculation was the proportion of the total tumour voxels predicted as 1p19q codeleted.

Three-class accuracies representing correct predictions for both IDH and 1p19q status were 90.4% on the NZ and 84.3% on the EGD testing dataset. Three-class AUC was 92.4% on the NZ and 91.2% on the EGD dataset. Input scores for the three-class AUC calculation were the relative proportion of predicted tumour voxels for each class. Table 3 depicts mean model performance over the NZ and EGD testing datasets.

**Table 3:** Model performance on NZ and EGD testing data.

|  | Metric | NZ | EGD |
| --- | --- | --- | --- |
| IDH | Accuracy | 93.3% ± 0.3 | 91.5% ± 0.1 |
| | Sensitivity | 69.8% ± 1.8 | 82.6% ± 1.4 |
| | Specificity | 97.7% ± 0.0 | 96.1% ± 0.6 |
| | AUC | 95.4% ± 0.2 | 95.8% ± 0.5 |
| 1p19q | Accuracy | 94.5% ± 0.7 | 87.5% ± 0.8 |
| | Sensitivity | 59.5% ± 4.1 | 54.0% ± 4.3 |
| | Specificity | 97.0% ± 0.8 | 94.4% ± 1.6 |
| | AUC | 92.5% ± 2.0 | 85.4% ± 0.4 |
| Three-Class | Accuracy | 90.4% ± 0.6 | 84.3% ± 0.2 |
| | AUC | 92.4% ± 1.3 | 91.2% ± 0.1 |

### IDH Confidence Score

The IDH confidence score is the proportion of tumour voxels predicted as IDH mutated (1p19q intact or codeleted). This score corresponds to the certainty of the model prediction, with low scores corresponding to certain predictions for IDH wild-type and high scores corresponding to confident IDH mutant predictions. Intermediate scores represent less certain classifications.

Analysis of predictive accuracy as a function of the confidence score demonstrates the expected increase in accuracy at each end of the confidence spectrum with greater than 95% accuracy over both datasets when the confidence score was less than 10% or greater than 90%. Observed predictions were less accurate for the intermediate confidence strata in each dataset with variation in the NZ dataset owing to the smaller sample size.

The model had high certainty for most predictions with confidence scores of less than 10% in 76.1% of cases for the NZ dataset and 54.1% in the EGD dataset. Corresponding values for confidence scores of greater than 90% were 6.7% and 15.1%. Table 4 summarises IDH predictive accuracy over each confidence score strata and their relative frequency in the NZ and EGD testing datasets.

**Table 4:**
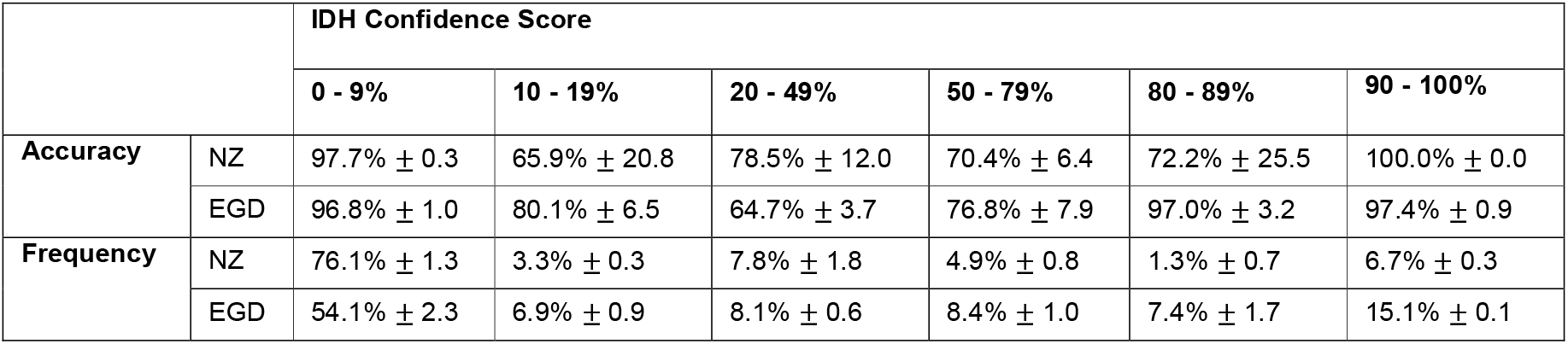
Accuracy and relative frequency across values of the IDH confidence score for the NZ and EGD testing datasets.

|  |  | IDH Confidence Score |  |  |  |  |  |
| --- | --- | --- | --- | --- | --- | --- | --- |
|  |  | 0 - 9% | 10 - 19% | 20 - 49% | 50 - 79% | 80 - 89% | 90 - 100% |
| **Accuracy** | NZ | 97.7% ± 0.3 | 65.9% ± 20.8 | 78.5% ± 12.0 | 70.4% ± 6.4 | 72.2% ± 25.5 | 100.0% ± 0.0 |
| | EGD | 96.8% ± 1.0 | 80.1% ± 6.5 | 64.7% ± 3.7 | 76.8% ± 7.9 | 97.0% ± 3.2 | 97.4% ± 0.9 |
| **Frequency** | NZ | 76.1% ± 1.3 | 3.3% ± 0.3 | 7.8% ± 1.8 | 4.9% ± 0.8 | 1.3% ± 0.7 | 6.7% ± 0.3 |
| | EGD | 54.1% ± 2.3 | 6.9% ± 0.9 | 8.1% ± 0.6 | 8.4% ± 1.0 | 7.4% ± 1.7 | 15.1% ± 0.1 |

## Discussion

In this study, a neural network method of IDH and 1p19q diagnosis was evaluated on a retrospective cohort of 205 patients with glioma treated in Auckland, New Zealand, and an independent cohort of 420 MR images from the Erasmus Glioma Database. Diagnostic accuracy on the NZ dataset for IDH was high at 93.3%, with very high specificity of 97.7%. 1p19q prediction was also accurate at 94.5% with specificity of 97.0%. Model performance remained high when evaluated on the external EGD data with accuracies of 91.5% and 87.5% for IDH and 1p19q, respectively. Specificity was also stable for both IDH and 1p19q with values of 96.1% and 94.4%, respectively.

Three-class classification accuracy represents the rate of correct prediction for both IDH and 1p19q status, and previous publications have assessed these markers separately [19,25]. Three-class accuracy was reduced compared to the separated IDH and 1p19q accuracies, which is expected given predictive errors in each category may occur in different subjects. A major advantage of the combined IDH and 1p19q approach presented here is this allows characterisation of diffuse gliomas into the WHO categories of GBM (IDH wild-type), astrocytoma (IDH mutated - 1p19q intact) and oligodendroglioma (IDH wild-type - 1p19q codeleted). While there are additional entity defining mutations in glioma, such as H3 k27-alteration in diffuse midline glioma, these tumours fit into the overarching categories and are less common, so IDH and 1p19q status allows categorisation of most tumours seen in clinical practice.

This is one of few studies to independently validate the use of neural networks for IDH prediction outside the TCIA dataset and is the largest combined testing dataset published to date. One of the major limitations of conducting research exclusively on TCIA data is the risk of poor model generalisation related to systematic patient and imaging differences between TCIA data and clinical datasets. Rates of IDH mutation are higher in TCIA data compared to longitudinal datasets where 20-30% of tumours are IDH mutated [9,32]. There are separate datasets for low- and high-grade glioma provided by the TCIA which may be the cause. Despite the demonstrated high internal performance on TCIA data, differences may mean these models fail to generalise. This was observed in reverse by Choi et al. who trained a model on regionally acquired data with a substantial fall in performance on TCIA images. Given the close correlation of IDH status with grade, there is a risk that models may be detecting non-target features related to systematic differences in the low-grade glioma and high-grade glioma TCIA datasets. This emphasises that caution should be taken in interpreting results where external validation has not been performed.

As a longitudinal cohort, the NZ dataset is representative of the kinds of cases which would be expected to be encountered in clinical practice. Rather than randomly allocating cases from the NZ cohort into training and testing datasets, the NZ cohort was split temporally, with the most recently treated patients forming the testing data. This mirrors a prospective approach, whereby a model trained in 2017 with the data available at that time, would be expected to perform in the same manner as the models presented here. Temporally splitting the dataset also reduces the likelihood of unseen confounding factors acting to artificially improve classification performance. This also indicates that the method is robust to changes in scan protocols over time, such as the increase in FLAIR volumetric imaging between the NZ training and testing datasets.

The TCIA, NZ and EGD training and testing datasets consist of a diverse range of imaging across multiple scan manufacturers, magnet strengths, and imaging protocols. Given the EGD data is completely independent of training data, the generalisation of model performance suggests that MRI-based IDH and 1p19q prediction would perform in a prospective clinical setting. Other similar approaches have utilised MRI sequences such as perfusion weighted imaging [47]. Radiologically, physiological sequences like diffusion and perfusion weighted imaging add crucial information to the evaluation of a tumour. From a model architecture perspective, additional sequences potentially introduce errors from misregistration and increase computational resource demands. Future work could investigate the value of additional sequences and this may increase performance. However, an advantage of this three-sequence approach is that it does not require additional scan time or alterations to MRI protocols.

The use of 2D vs 3D computational methods in MRI involves trade-offs related to 2D acquisition and anisotropy of spatial resolution [48]. One of the key considerations is that MRI can be acquired slice-wise or volumetrically. In both TCIA and NZ datasets, volumetric imaging was performed mainly for T1 post-contrast sequences. Volumetric FLAIR sequences were occasionally performed in the NZ dataset and volumetric T2 sequences were not present. 3D reconstruction of non-volumetric imaging results in interpolation between slices. When the interpolated data is fed into a 3D network, the spatial resolution in the interpolated axis is reduced, so the model receives less information in this dimension compared to the in-plane axes. The 3D T2-net trained by Yogananda et al. utilised TCIA T2 imaging exclusively, however volumetric T2 imaging was not performed in the TCIA dataset with a mean slice thickness of 4.5 mm. This implies a high level of redundancy in the out-of-plane axis of their 3D model. Interpolation between slices is less of a consideration using a 2D approach and there are considerable computational advantages of 2D models from a resource perspective. This allows the utilisation of larger image patches which present more of the brain to the model at each iteration, potentially allowing anatomical contextualisation of the tumour relative to normal brain structures. This motivated the decision to use a 2D method, although volumetric imaging will likely become more widespread and advances in computational performance will alleviate limitations on the model architecture.

Data leakage is a recognised issue, whereby performance is artificially increased due to cross-over in data between training and testing datasets [20]. This typically occurs when different MRI slices from the same patient are split between training and testing datasets, which has been demonstrated to increase the classification accuracy by 6% [20]. The use of transfer learning has the potential to introduce a different form of data leak if pre-trained models are repurposed for new classification problems on the same data using different cross-validation allocations [19,25,26]. In this case where both 1p19q and MGMT status are associated with IDH status, the use of transfer learning from a pre-trained IDH classification model may have artificially increased accuracy. This represents a potentially unrecognised form of data leakage which should be acknowledged in experimental design and is avoided by our combined approach.

Our method utilises automated segmentation to generate the training input segmentations. Manual segmentation is highly time consuming and requires expert radiological knowledge which can be averted by automated segmentation. Although manual segmentation is the gold standard, well-trained automated techniques approach the level of interobserver variability [49]. The high performance of the IDH and 1p19q predictive model in our case validates the use of automated segmentation during training, even if the individual segmentations have not been verified by a radiologist. The primary endpoint here is IDH and 1p19q prediction, and while poor quality input segmentations during training are likely to degrade predictive accuracy, the high performance of the final model means the training segmentations must have been satisfactory.

An advantage of the segmentation-classification method is the output segmentation can be visually assessed. This is a possible quality control step and provides reassurance that the model is making IDH and 1p19q predictions based on tumour related features. Examining the segmentations may also be useful for investigating the features the model is using to make classifications. A common critique of AI approaches in medicine is that these methods remain largely inexplicable. The segmentation approach adopted in this work has the potential to address some of these concerns. Like others who have used this method, we do not claim that heterogeneity in the IDH and 1p19q segmentation is related to variability in the genetic composition of the analysed tumour [19]. This approach is likely applicable in other aspects of radiology, such as paediatric brain tumours where there are several important molecular subtypes.

A limitation of this study is the relatively low sensitivity which reflects missed cases of IDH mutation and 1p19q codeletion. This is countered by the very high specificity for both markers, resulting in an overall high accuracy. While descriptive statistics like sensitivity and specificity are crucial to the evaluation of a diagnostic test, the clinician must treat the patient in front of them. Knowledge of whether a particular result can be trusted is an important part of clinical reasoning, and an equivocal result warrants increased skepticism. The IDH confidence score is useful in this respect, and we have demonstrated a relationship between confidence score and accuracy, with greater than 95% accuracy with scores at either end of the confidence spectrum. Importantly, most individual confidence scores were <10% or >90% indicating that most results provide useful information. A metric of diagnostic certainty allows clinicians to exercise judgement and integrate results with other clinical and radiological information about individual patients.

From a radiological perspective, one of the major appeals of this method is that it provides information which is not readily available from visual assessment. While we have demonstrated our model can distinguish histological subtypes of diffuse glioma, its application depends on sound judgement to ensure that it is used in appropriate clinical settings. Uncommon entities such as tumefactive demyelination may mimic tumours to the untrained eye and result in errors if algorithms such as this are used outside their intended scope. This is a limitation of AI methods throughout medicine and emphasises the importance of expert judgement so that they are applied appropriately.

The major impact is the possibility of reliable IDH and 1p19q diagnosis before surgery by imaging which is already performed as the standard of care in glioma. This method is fast and increases the diagnostic utility of MRI without the need for changes to imaging protocols or manual input. One of the limitations of pathological molecular diagnosis is the turnaround time for results, which translates to a delay in post-surgical treatment decisions – as well as an anxious wait for patients and their families.

Our method can provide molecular information prior to resection which could inform decisions about surgery and other pre-operative treatments. There are several promising IDH specific treatment agents including IDH inhibitors [50]. Non-invasive IDH diagnosis offers the possibility of selecting patients for IDH dependent therapy before biopsy or surgery. This is likely to be a highly useful application if IDH specific therapies are proven to be beneficial, especially in the case of neoadjuvant treatments. Preoperative molecular diagnosis may assist surgical planning and allow neurosurgeons to optimise the resection according to mutation status. A survival benefit has been demonstrated for the complete resection of both enhancing and non-enhancing tissue in IDH mutated astrocytoma [28]. Given more extensive resection carries an increased risk, this approach may only be justified in IDH mutated tumours as these patients may live for decades if there is no residual tumour [28]. Accurate pre-surgical IDH diagnosis may facilitate patient selection for extensive resection as well as allow better patient information of the potential benefits of surgery. High IDH specificity is desirable in both these applications to prevent patients with IDH wild-type tumours being submitted to additional risks without the desired benefit.

## Conclusion

High IDH and 1p19q classification accuracies were achieved on a retrospective cohort of 205 patients with diffuse glioma treated at Auckland City Hospital, New Zealand. Performance remained high when assessed on an external cohort of 420 scans from the EGD demonstrating the generalisability of this method. The ability to non-invasively characterise tumours with routine MRI sequences according to WHO 2021 criteria demonstrates the potential of this as a useful tool in glioma diagnosis with implications for treatment.

## Data Availability

MRI data from the Erasmus Glioma Database is available with instructions at the DOI 10.1016/j.dib.2021.107191. TCIA data is available from https://doi.org/10.7937/K9/TCIA.2016.RNYFUYE9 and http://doi.org/10.7937/K9/TCIA.2016.L4LTD3TK. Data from the NZ dataset is not publicly available.

## Acknowledgements

Akul Sharma and Toby Struthers. Study participants. EGD and TCIA contributors. Organizers of BraTS. Neurosurgery Department at Auckland City Hospital. Radiology Departments at Auckland City, Middlemore and North Shore Hospitals.

## References

1. Ostrum QT, Gittleman H, Fulop J, et al. CBTRUS statistical report: primary brain and central nervous system tumors diagnosed in the United States in 2008–2012.Neuro Oncol. 2015;17(suppl 4): v1–v62.

2. Yan H, Parsons DW, Jin G, et al. IDH1 and IDH2 Mutations in Gliomas. New England Journal of Medicine. 2009;360(8):765–773.

3. Garton ALA, Kinslow CJ, Rae AI, et al. Extent of resection, molecular signature, and survival in 1p19q-codeleted gliomas. Journal of Neurosurgery JNS. 2021;134(5):1357 – 1367.

4. Parsons DW, Jones S, Zhang X, et al. An integrated genomic analysis of human glioblastoma multiforme. Science. 2008;321(5897):1807–1812.

5. Louis DN, Perry A, Reifenberger G, et al. The 2016 World Health Organization classification of tumors of the central nervous system: a summary. Acta neuropathologica. 2016;131(6):803–820.

6. Louis DN, Perry A, Wesseling P, et al. The 2021 WHO classification of tumors of the central nervous system: a summary. Neuro-oncology.2021;23(8):1231–1251.

7. Puduvalli VK, Hashmi M, McAllister LD, et al. Anaplastic oligodendrogliomas: prognostic factors for tumor recurrence and survival. Oncology. 2003;65(3):259–266.

8. Deng L, Xiong P, Luo Y, et al. Association between IDH1/2 mutations and brain glioma grade. Oncology letters. 2018;16(4):5405–5409.

9. Choi YS, Bae S, Chang JH, et al. Fully automated hybrid approach to predict the IDH mutation status of gliomas via deep learning and radiomics. Neuro-oncology. 2021;23(2):304–313.

10. Forst DA, Nahed BV, Loeffler JS, Batchelor TT. Low-grade gliomas. The oncologist. 2014;19(4):403.

11. Rasmussen BK, Hansen S, Laursen RJ, et al. Epidemiology of glioma: clinical characteristics, symptoms, and predictors of glioma patients grade I–IV in the Danish Neuro-Oncology Registry. Journal of Neuro-oncology. 2017;135(3):571–579.

12. Nagashima H, Tanaka K, Sasayama T, et al. Diagnostic value of glutamate with 2-hydroxyglutarate in magnetic resonance spectroscopy for IDH1 mutant glioma. Neurooncology. 2016;18(11):1559–1568.

13. Lombardi G, Corona G, Bellu L, et al. Diagnostic value of plasma and urinary 2-hydroxyglutarate to identify patients with isocitrate dehydrogenase-mutated glioma. The Oncologist. 2015;20(5):562.

14. Miller AM, Shah RH, Pentsova EI, et al. Tracking tumour evolution in glioma through liquid biopsies of cerebrospinal fluid. Nature. 2019;565(7741):654–658.

15. Corell A, Ferreyra VS, Hoefling N, et al. The clinical significance of the T2-FLAIR mismatch sign in grade II and III gliomas: a population-based study. BMC cancer. 2020; 20:1–10.

16. Deguchi S, Oishi T, Mitsuya K, et al. Clinicopathological analysis of T2-FLAIR mismatch sign in lower-grade gliomas. Scientific reports. 2020;10(1):1–6.

17. Smits M. Imaging of oligodendroglioma. The British Journal of Radiology. 2016;89(1060):20150857.

18. Ellingson BM, Bendszus M, Boxerman J, et al. Consensus recommendations for a standardized brain tumor imaging protocol in clinical trials. Neuro-oncology. 2015;17(9):1188–1198.

19. Yogananda CGB, Shah BR, Vejdani-Jahromi M, et al. A novel fully automated MRI-based deep-learning method for classification of IDH mutation status in brain gliomas. Neuro-Oncology. 2019;22(3):402–411.

20. Nalawade S, Murugesan GK, Vejdani-Jahromi M, et al. Classification of brain tumor isocitrate dehydrogenase status using MRI and deep learning. Journal of Medical Imaging. 2019;6(4):046003.

21. Li Z, Wang Y, Yu J, Guo Y, Cao W. Deep learning based radiomics (DLR) and its usage in noninvasive IDH1 prediction for low grade glioma. Scientific reports. 2017;7(1):1–11.

22. Chang P, Grinband J, Weinberg BD, et al. Deep-learning convolutional neural networks accurately classify genetic mutations in gliomas. American Journal of Neuroradiology. 2018;39(7):1201–1207.

23. Chang K, Harrison XB, Zhou H, et al. Residual convolutional neural network for the determination of IDH status in low-and high-grade gliomas from MR imaging. Clinical Cancer Research. 2018;24(5):1073–1081.

24. Liang S, Zhang R, Liang D, et al. Multimodal 3D DenseNet for IDH genotype prediction in gliomas. Genes. 2018;9(8):382.

25. Yogananda CGB, Shah BR, Frank FY, et al. A novel fully automated MRI-based deep-learning method for classification of 1p/19q co-deletion status in brain gliomas. Neuro-oncology advances. 2020;2

26. Yogananda CGB, Shah BR, Nalawade SS, et al. MRI-Based Deep-Learning Method for Determining Glioma MGMT Promoter Methylation Status. American Journal of Neuroradiology. 2021;42(5):845–852.

27. Clark K, Vendt B, Smith K, et al. The Cancer Imaging Archive (TCIA): maintaining and operating a public information repository. Journal of digital imaging. 2013;26(6):1045–1057.

28. Beiko J, Suki D, Hess KR, et al. IDH1 mutant malignant astrocytomas are more amenable to surgical resection and have a survival benefit associated with maximal surgical resection. Neuro-oncology. 2014;16(1):81–91.

29. Jacobo JA, Perez SM, Moreno-Jimenez S. The Role of Neoadjuvant Therapy to Improve the Extent of Resection in “Unresectable” Gliomas. World Neurosurgery. 2020.

30. Scarpace L, Mikkelsen T, Cha S, et al. Radiology Data from The Cancer Genome Atlas Glioblastoma Multiforme [TCGA-GBM] collection. The Cancer Imaging Archive. 2016.

31. Pedano N, Flanders AE, Scarpace L, et al. Radiology Data from The Cancer Genome Atlas Low Grade Glioma [TCGA-LGG] collection. The Cancer Imaging Archive. 2016.

32. Voort SR, Incekara F, Wijnenga MMJ, et al. The Erasmus Glioma Database (EGD): Structural MRI scans, WHO 2016 subtypes, and segmentations of 774 patients with glioma. Data in brief. 2021; 37:107191.

33. Li X, Morgan PS, Ashburner J, Smith J, Rorden C. The first step for neuroimaging data analysis: DICOM to NIfTI conversion. Journal of neuroscience methods. 2016; 264:47–56.

34. Beare R, Lowekamp B, Yaniv Z. Image Segmentation, Registration and Characterization in R with SimpleITK. Journal of Statistical Software. 2018;86(8):1–35.

35. Yaniv Z, Lowekamp BC, Johnson HJ, Beare R. SimpleITK Image-Analysis Notebooks: a Collaborative Environment for Education and Reproducible Research. Journal of Digital Imaging. 2018;31(3):290–303.

36. Lowekamp BC, Chen DT, Ibáñez L, Blezek D. The Design of SimpleITK. Frontiers in neuroinformatics. 2013; 7:45.

37. Rohlfing T, Zahr NM, Sullivan EV, Pfefferbaum A. The SRI24 multichannel atlas of normal adult human brain structure. Human brain mapping. 2010;31(5):798–819.

38. Baid U, Ghodasara S, Mohan S, et al. The RSNA-ASNR-MICCAI BraTS 2021 Benchmark on Brain Tumor Segmentation and Radiogenomic Classification. 2021.

39. Menze BH, Jakab A, Bauer S, et al. The Multimodal Brain Tumor Image Segmentation Benchmark (BRATS). IEEE transactions on medical imaging. 2015;34(10):1993–2024.

40. Bakas S, Akbari H, Sotiras A, et al. Advancing the cancer genome atlas glioma MRI collections with expert segmentation labels and radiomic features. Scientific data. 2017;4(1):1–13.

41. Fonov V, Evans AC., Botteron K, Almli CR, McKinstry RC, Collins DL. Unbiased average age-appropriate atlases for pediatric studies. NeuroImage. 2011;54(1):313–327.

42. Fonov VS, Evans AC, McKinstry RC, Almli CR, Collins DL. Unbiased nonlinear average age-appropriate brain templates from birth to adulthood. NeuroImage. 2009; 47:S102. Organization for Human Brain Mapping 2009 Annual Meeting.

43. Abadi M, Agarwal A, Barham P, et al. Tensorflow: Large-scale machine learning on heterogeneous distributed systems. arXiv preprint arXiv:1603.04467.2016.

44. Ronneberger O, Fischer P, Brox T. U-net: Convolutional networks for biomedical image segmentation. In: 234–241 Springer; 2015.

45. Huang G, Liu Z, Van Der Maaten L, Weinberger KQ. Densely connected convolutional networks. In: 4700–4708; 2017.

46. Pedregosa F, Varoquaux G, Gramfort A, et al. Scikit-learn: Machine Learning in Python.Journal of Machine Learning Research. 2011; 12:2825–2830.

47. Choi KS, Choi SH, Jeong B. Prediction of IDH genotype in gliomas with dynamic susceptibility contrast perfusion MR imaging using an explainable recurrent neural network. Neuro-oncology. 2019;21(9):1197–1209.

48. McHugh H, Talou GM, Wang A. 2D Dense-UNet: A Clinically Valid Approach to Automated Glioma Segmentation. In: 69–80Springer; 2020.

49. Rudie JD, Weiss DA, Saluja R, et al. Multi-disease segmentation of gliomas and white matter hyperintensities in the BraTS data using a 3D convolutional neural network. Frontiers in computational neuroscience. 2019; 13:84.

50. Mellinghoff IK, Van Den Bent MJ, Clarke JL, et al. INDIGO: A global, randomized, double-blind, phase III study of vorasidenib (VOR; AG-881) vs placebo in patients (pts) with residual or recurrent grade II glioma with an isocitrate dehydrogenase 1/2 (IDH1/2) mutation. Journal of Clinical Oncology. 2020;38(15_suppl)

