## Supplemental Data for "IDH and 1p19q Diagnosis in Diffuse Glioma from Preoperative MRI Using Artificial Intelligence"

### Additional Figures

**Table 1:** Confusion matrix of performance on the NZ testing dataset in experiment one.

| **NZ Experiment One** | **Actual** | | |
| --- | --- | --- | --- |
| **Predicted** | IDH Wild-Type | IDH Mutated - 1p19q Intact | IDH Mutated - 1p19q Codeleted |
| IDH Wild-Type | 169 | 6 | 4 |
| IDH Mutated - 1p19q Intact | 3 | 9 | 2 |
| IDH Mutated - 1p19q Codeleted | 1 | 3 | 8 |

**Table 2:** Confusion matrix of performance on the NZ testing dataset in experiment two.

| **NZ Experiment Two** | **Actual** | | |
| --- | --- | --- | --- |
| **Predicted** | IDH Wild-Type | IDH Mutated - 1p19q Intact | IDH Mutated - 1p19q Codeleted |
| IDH Wild-Type | 169 | 5 | 4 |
| IDH Mutated - 1p19q Intact | 3 | 8 | 1 |
| IDH Mutated - 1p19q Codeleted | 1 | 5 | 9 |

**Table 3:**  Confusion matrix of performance on the NZ testing dataset in experiment three.

| **NZ Experiment Three** | **Actual** | | |
| --- | --- | --- | --- |
| **Predicted** | IDH Wild-Type | IDH Mutated - 1p19q Intact | IDH Mutated - 1p19q Codeleted |
| IDH Wild-Type | 169 | 5 | 5 |
| IDH Mutated - 1p19q Intact | 3 | 7 | 1 |
| IDH Mutated - 1p19q Codeleted | 1 | 6 | 8 |

**Table 4:** Confusion matrix of performance on the EGD testing dataset in experiment one.

| **EGD Experiment One** | **Actual** | | |
| --- | --- | --- | --- |
| **Predicted** | IDH Wild-Type | IDH Mutated - 1p19q Intact | IDH Mutated - 1p19q Codeleted |
| IDH Wild-Type | 265 | 7 | 18 |
| IDH Mutated - 1p19q Intact | 8 | 54 | 18 |
| IDH Mutated - 1p19q Codeleted | 3 | 12 | 35 |

**Table 5:** Confusion matrix of performance on the EGD testing dataset in experiment two.

| **EGD Experiment Two** | **Actual** | | |
| --- | --- | --- | --- |
| **Predicted** | IDH Wild-Type | IDH Mutated - 1p19q Intact | IDH Mutated - 1p19q Codeleted |
| IDH Wild-Type | 267 | 9 | 18 |
| IDH Mutated - 1p19q Intact | 6 | 49 | 14 |
| IDH Mutated - 1p19q Codeleted | 3 | 15 | 39 |

**Table 6:** Confusion matrix of performance on the EGD testing dataset in experiment three.

| **EGD Experiment Three** | **Actual** | | |
| --- | --- | --- | --- |
| **Predicted** | IDH Wild-Type | IDH Mutated - 1p19q Intact | IDH Mutated - 1p19q Codeleted |
| IDH Wild-Type | 264 | 6 | 17 |
| IDH Mutated - 1p19q Intact | 5 | 48 | 13 |
| IDH Mutated - 1p19q Codeleted | 7 | 19 | 41 |

**Table 7:** Labels, ages, IDH and 1p19q status of the cases evaluated in the EGD testing dataset.

| **Subject** | **IDH** | **1p19q** |
| --- | --- | --- |
| EGD-0004 | Mutated | Codeleted |
| EGD-0008 | Mutated | Codeleted |
| EGD-0009 | Wild-Type | - |
| EGD-0011 | Mutated | Codeleted |
| EGD-0014 | Wild-Type | - |
| EGD-0015 | Wild-Type | - |
| EGD-0020 | Mutated | Codeleted |
| EGD-0022 | Mutated | Codeleted |
| EGD-0024 | Mutated | Intact |
| EGD-0026 | Wild-Type | - |
| EGD-0029 | Mutated | Intact |
| EGD-0031 | Wild-Type | - |
| EGD-0033 | Mutated | Codeleted |
| EGD-0035 | Wild-Type | - |
| EGD-0041 | Wild-Type | - |
| EGD-0045 | Wild-Type | - |
| EGD-0047 | Mutated | Intact |
| EGD-0050 | Wild-Type | - |
| EGD-0052 | Wild-Type | - |
| EGD-0053 | Wild-Type | - |
| EGD-0055 | Wild-Type | - |
| EGD-0057 | Mutated | Codeleted |
| EGD-0058 | Wild-Type | - |
| EGD-0064 | Wild-Type | - |
| EGD-0066 | Mutated | Codeleted |
| EGD-0068 | Mutated | Codeleted |
| EGD-0072 | Mutated | Intact |
| EGD-0073 | Wild-Type | - |
| EGD-0074 | Mutated | Intact |
| EGD-0075 | Wild-Type | - |
| EGD-0080 | Wild-Type | - |
| EGD-0081 | Wild-Type | - |
| EGD-0083 | Wild-Type | - |
| EGD-0085 | Wild-Type | - |
| EGD-0086 | Mutated | Intact |
| EGD-0088 | Wild-Type | - |
| EGD-0089 | Wild-Type | - |
| EGD-0090 | Mutated | Intact |
| EGD-0094 | Wild-Type | - |
| EGD-0097 | Wild-Type | - |
| EGD-0099 | Wild-Type | - |
| EGD-0100 | Wild-Type | - |
| EGD-0101 | Mutated | Intact |
| EGD-0102 | Wild-Type | - |
| EGD-0105 | Wild-Type | - |
| EGD-0106 | Mutated | Intact |
| EGD-0107 | Mutated | Intact |
| EGD-0109 | Wild-Type | - |
| EGD-0110 | Wild-Type | - |
| EGD-0111 | Wild-Type | - |
| EGD-0112 | Wild-Type | - |
| EGD-0114 | Wild-Type | - |
| EGD-0115 | Wild-Type | - |
| EGD-0118 | Mutated | Intact |
| EGD-0121 | Wild-Type | - |
| EGD-0122 | Mutated | Intact |
| EGD-0123 | Mutated | Codeleted |
| EGD-0126 | Wild-Type | - |
| EGD-0127 | Wild-Type | - |
| EGD-0128 | Mutated | Intact |
| EGD-0131 | Wild-Type | - |
| EGD-0132 | Wild-Type | - |
| EGD-0133 | Mutated | Codeleted |
| EGD-0134 | Wild-Type | - |
| EGD-0141 | Wild-Type | - |
| EGD-0143 | Mutated | Codeleted |
| EGD-0144 | Wild-Type | - |
| EGD-0145 | Mutated | Intact |
| EGD-0146 | Mutated | Codeleted |
| EGD-0152 | Wild-Type | - |
| EGD-0155 | Wild-Type | - |
| EGD-0156 | Wild-Type | - |
| EGD-0158 | Wild-Type | - |
| EGD-0160 | Mutated | Intact |
| EGD-0161 | Wild-Type | - |
| EGD-0162 | Mutated | Codeleted |
| EGD-0165 | Wild-Type | - |
| EGD-0166 | Wild-Type | - |
| EGD-0167 | Wild-Type | - |
| EGD-0168 | Wild-Type | - |
| EGD-0171 | Wild-Type | - |
| EGD-0172 | Wild-Type | - |
| EGD-0176 | Wild-Type | - |
| EGD-0177 | Mutated | Codeleted |
| EGD-0179 | Mutated | Codeleted |
| EGD-0182 | Wild-Type | - |
| EGD-0183 | Wild-Type | - |
| EGD-0185 | Mutated | Intact |
| EGD-0186 | Mutated | Codeleted |
| EGD-0187 | Mutated | Codeleted |
| EGD-0188 | Mutated | Intact |
| EGD-0191 | Mutated | Intact |
| EGD-0192 | Wild-Type | - |
| EGD-0193 | Wild-Type | - |
| EGD-0194 | Mutated | Codeleted |
| EGD-0196 | Wild-Type | - |
| EGD-0197 | Wild-Type | - |
| EGD-0198 | Wild-Type | - |
| EGD-0199 | Mutated | Codeleted |
| EGD-0202 | Wild-Type | - |
| EGD-0203 | Mutated | Intact |
| EGD-0204 | Wild-Type | - |
| EGD-0206 | Wild-Type | - |
| EGD-0208 | Mutated | Intact |
| EGD-0211 | Mutated | Intact |
| EGD-0213 | Wild-Type | - |
| EGD-0214 | Mutated | Codeleted |
| EGD-0215 | Wild-Type | - |
| EGD-0216 | Wild-Type | - |
| EGD-0217 | Mutated | Codeleted |
| EGD-0218 | Wild-Type | - |
| EGD-0219 | Wild-Type | - |
| EGD-0221 | Mutated | Codeleted |
| EGD-0222 | Wild-Type | - |
| EGD-0223 | Wild-Type | - |
| EGD-0224 | Wild-Type | - |
| EGD-0226 | Wild-Type | - |
| EGD-0227 | Mutated | Intact |
| EGD-0228 | Mutated | Codeleted |
| EGD-0229 | Mutated | Intact |
| EGD-0232 | Wild-Type | - |
| EGD-0233 | Mutated | Codeleted |
| EGD-0234 | Wild-Type | - |
| EGD-0236 | Mutated | Intact |
| EGD-0237 | Wild-Type | - |
| EGD-0238 | Wild-Type | - |
| EGD-0240 | Mutated | Intact |
| EGD-0241 | Mutated | Intact |
| EGD-0242 | Wild-Type | - |
| EGD-0243 | Mutated | Intact |
| EGD-0244 | Mutated | Codeleted |
| EGD-0247 | Wild-Type | - |
| EGD-0250 | Wild-Type | - |
| EGD-0253 | Mutated | Codeleted |
| EGD-0254 | Wild-Type | - |
| EGD-0255 | Wild-Type | - |
| EGD-0256 | Wild-Type | - |
| EGD-0262 | Wild-Type | - |
| EGD-0263 | Wild-Type | - |
| EGD-0264 | Wild-Type | - |
| EGD-0268 | Mutated | Intact |
| EGD-0270 | Wild-Type | - |
| EGD-0275 | Wild-Type | - |
| EGD-0280 | Wild-Type | - |
| EGD-0281 | Wild-Type | - |
| EGD-0282 | Wild-Type | - |
| EGD-0285 | Wild-Type | - |
| EGD-0286 | Mutated | Codeleted |
| EGD-0290 | Wild-Type | - |
| EGD-0292 | Wild-Type | - |
| EGD-0293 | Wild-Type | - |
| EGD-0294 | Wild-Type | - |
| EGD-0297 | Mutated | Intact |
| EGD-0298 | Wild-Type | - |
| EGD-0299 | Wild-Type | - |
| EGD-0301 | Mutated | Intact |
| EGD-0302 | Wild-Type | - |
| EGD-0303 | Wild-Type | - |
| EGD-0305 | Wild-Type | - |
| EGD-0306 | Mutated | Intact |
| EGD-0309 | Mutated | Intact |
| EGD-0311 | Wild-Type | - |
| EGD-0312 | Mutated | Codeleted |
| EGD-0313 | Wild-Type | - |
| EGD-0314 | Wild-Type | - |
| EGD-0315 | Wild-Type | - |
| EGD-0318 | Wild-Type | - |
| EGD-0319 | Wild-Type | - |
| EGD-0321 | Wild-Type | - |
| EGD-0324 | Wild-Type | - |
| EGD-0325 | Wild-Type | - |
| EGD-0328 | Wild-Type | - |
| EGD-0330 | Mutated | Codeleted |
| EGD-0331 | Wild-Type | - |
| EGD-0332 | Mutated | Codeleted |
| EGD-0334 | Wild-Type | - |
| EGD-0336 | Wild-Type | - |
| EGD-0337 | Wild-Type | - |
| EGD-0338 | Wild-Type | - |
| EGD-0340 | Mutated | Intact |
| EGD-0342 | Wild-Type | - |
| EGD-0343 | Wild-Type | - |
| EGD-0344 | Wild-Type | - |
| EGD-0345 | Wild-Type | - |
| EGD-0346 | Mutated | Intact |
| EGD-0349 | Wild-Type | - |
| EGD-0351 | Mutated | Codeleted |
| EGD-0352 | Mutated | Codeleted |
| EGD-0353 | Wild-Type | - |
| EGD-0354 | Wild-Type | - |
| EGD-0355 | Mutated | Intact |
| EGD-0356 | Mutated | Codeleted |
| EGD-0357 | Wild-Type | - |
| EGD-0358 | Wild-Type | - |
| EGD-0363 | Wild-Type | - |
| EGD-0365 | Wild-Type | - |
| EGD-0366 | Wild-Type | - |
| EGD-0367 | Wild-Type | - |
| EGD-0368 | Mutated | Intact |
| EGD-0369 | Mutated | Codeleted |
| EGD-0370 | Wild-Type | - |
| EGD-0375 | Wild-Type | - |
| EGD-0376 | Wild-Type | - |
| EGD-0377 | Mutated | Intact |
| EGD-0378 | Mutated | Intact |
| EGD-0380 | Wild-Type | - |
| EGD-0381 | Wild-Type | - |
| EGD-0382 | Wild-Type | - |
| EGD-0383 | Wild-Type | - |
| EGD-0385 | Mutated | Intact |
| EGD-0387 | Mutated | Intact |
| EGD-0388 | Wild-Type | - |
| EGD-0389 | Wild-Type | - |
| EGD-0390 | Wild-Type | - |
| EGD-0391 | Wild-Type | - |
| EGD-0392 | Wild-Type | - |
| EGD-0394 | Wild-Type | - |
| EGD-0395 | Wild-Type | - |
| EGD-0397 | Wild-Type | - |
| EGD-0400 | Wild-Type | - |
| EGD-0401 | Wild-Type | - |
| EGD-0402 | Mutated | Codeleted |
| EGD-0403 | Mutated | Codeleted |
| EGD-0405 | Wild-Type | - |
| EGD-0407 | Wild-Type | - |
| EGD-0410 | Mutated | Intact |
| EGD-0411 | Mutated | Codeleted |
| EGD-0413 | Mutated | Intact |
| EGD-0414 | Wild-Type | - |
| EGD-0415 | Mutated | Intact |
| EGD-0416 | Wild-Type | - |
| EGD-0417 | Wild-Type | - |
| EGD-0418 | Wild-Type | - |
| EGD-0420 | Mutated | Codeleted |
| EGD-0421 | Mutated | Codeleted |
| EGD-0423 | Wild-Type | - |
| EGD-0424 | Wild-Type | - |
| EGD-0425 | Wild-Type | - |
| EGD-0426 | Mutated | Codeleted |
| EGD-0427 | Wild-Type | - |
| EGD-0428 | Wild-Type | - |
| EGD-0431 | Wild-Type | - |
| EGD-0432 | Wild-Type | - |
| EGD-0433 | Wild-Type | - |
| EGD-0434 | Mutated | Codeleted |
| EGD-0438 | Wild-Type | - |
| EGD-0439 | Mutated | Intact |
| EGD-0440 | Wild-Type | - |
| EGD-0442 | Wild-Type | - |
| EGD-0443 | Wild-Type | - |
| EGD-0447 | Wild-Type | - |
| EGD-0448 | Wild-Type | - |
| EGD-0449 | Wild-Type | - |
| EGD-0450 | Mutated | Codeleted |
| EGD-0451 | Wild-Type | - |
| EGD-0452 | Wild-Type | - |
| EGD-0453 | Mutated | Intact |
| EGD-0455 | Wild-Type | - |
| EGD-0457 | Wild-Type | - |
| EGD-0461 | Mutated | Codeleted |
| EGD-0463 | Wild-Type | - |
| EGD-0464 | Wild-Type | - |
| EGD-0466 | Mutated | Intact |
| EGD-0469 | Wild-Type | - |
| EGD-0470 | Wild-Type | - |
| EGD-0472 | Mutated | Codeleted |
| EGD-0474 | Mutated | Codeleted |
| EGD-0479 | Wild-Type | - |
| EGD-0480 | Wild-Type | - |
| EGD-0482 | Wild-Type | - |
| EGD-0485 | Wild-Type | - |
| EGD-0487 | Mutated | Codeleted |
| EGD-0488 | Wild-Type | - |
| EGD-0490 | Mutated | Codeleted |
| EGD-0492 | Mutated | Intact |
| EGD-0494 | Wild-Type | - |
| EGD-0495 | Mutated | Codeleted |
| EGD-0499 | Mutated | Codeleted |
| EGD-0501 | Wild-Type | - |
| EGD-0502 | Wild-Type | - |
| EGD-0503 | Wild-Type | - |
| EGD-0505 | Wild-Type | - |
| EGD-0506 | Wild-Type | - |
| EGD-0507 | Wild-Type | - |
| EGD-0508 | Wild-Type | - |
| EGD-0509 | Mutated | Codeleted |
| EGD-0511 | Mutated | Intact |
| EGD-0512 | Wild-Type | - |
| EGD-0514 | Mutated | Codeleted |
| EGD-0515 | Wild-Type | - |
| EGD-0516 | Wild-Type | - |
| EGD-0518 | Mutated | Codeleted |
| EGD-0521 | Mutated | Intact |
| EGD-0523 | Mutated | Intact |
| EGD-0526 | Wild-Type | - |
| EGD-0529 | Wild-Type | - |
| EGD-0530 | Mutated | Intact |
| EGD-0532 | Wild-Type | - |
| EGD-0535 | Wild-Type | - |
| EGD-0536 | Mutated | Intact |
| EGD-0537 | Mutated | Codeleted |
| EGD-0539 | Wild-Type | - |
| EGD-0541 | Mutated | Codeleted |
| EGD-0544 | Wild-Type | - |
| EGD-0545 | Wild-Type | - |
| EGD-0547 | Wild-Type | - |
| EGD-0552 | Mutated | Codeleted |
| EGD-0553 | Mutated | Intact |
| EGD-0558 | Wild-Type | - |
| EGD-0561 | Mutated | Codeleted |
| EGD-0563 | Mutated | Intact |
| EGD-0564 | Mutated | Intact |
| EGD-0565 | Mutated | Intact |
| EGD-0567 | Mutated | Intact |
| EGD-0568 | Mutated | Intact |
| EGD-0572 | Mutated | Codeleted |
| EGD-0575 | Wild-Type | - |
| EGD-0577 | Mutated | Intact |
| EGD-0579 | Wild-Type | - |
| EGD-0580 | Wild-Type | - |
| EGD-0581 | Wild-Type | - |
| EGD-0585 | Mutated | Intact |
| EGD-0586 | Mutated | Intact |
| EGD-0587 | Wild-Type | - |
| EGD-0589 | Wild-Type | - |
| EGD-0591 | Wild-Type | - |
| EGD-0592 | Wild-Type | - |
| EGD-0593 | Wild-Type | - |
| EGD-0594 | Wild-Type | - |
| EGD-0595 | Wild-Type | - |
| EGD-0596 | Wild-Type | - |
| EGD-0598 | Mutated | Codeleted |
| EGD-0599 | Wild-Type | - |
| EGD-0600 | Wild-Type | - |
| EGD-0601 | Mutated | Codeleted |
| EGD-0602 | Wild-Type | - |
| EGD-0603 | Wild-Type | - |
| EGD-0604 | Wild-Type | - |
| EGD-0605 | Wild-Type | - |
| EGD-0606 | Mutated | Codeleted |
| EGD-0608 | Wild-Type | - |
| EGD-0611 | Wild-Type | - |
| EGD-0612 | Wild-Type | - |
| EGD-0617 | Wild-Type | - |
| EGD-0621 | Wild-Type | - |
| EGD-0622 | Wild-Type | - |
| EGD-0624 | Wild-Type | - |
| EGD-0625 | Wild-Type | - |
| EGD-0626 | Mutated | Intact |
| EGD-0627 | Wild-Type | - |
| EGD-0629 | Wild-Type | - |
| EGD-0631 | Mutated | Codeleted |
| EGD-0632 | Wild-Type | - |
| EGD-0633 | Mutated | Codeleted |
| EGD-0635 | Wild-Type | - |
| EGD-0636 | Mutated | Codeleted |
| EGD-0638 | Mutated | Codeleted |
| EGD-0639 | Wild-Type | - |
| EGD-0641 | Wild-Type | - |
| EGD-0643 | Wild-Type | - |
| EGD-0647 | Mutated | Intact |
| EGD-0648 | Wild-Type | - |
| EGD-0655 | Mutated | Intact |
| EGD-0656 | Mutated | Codeleted |
| EGD-0657 | Wild-Type | - |
| EGD-0658 | Wild-Type | - |
| EGD-0659 | Wild-Type | - |
| EGD-0661 | Wild-Type | - |
| EGD-0664 | Wild-Type | - |
| EGD-0665 | Wild-Type | - |
| EGD-0667 | Wild-Type | - |
| EGD-0668 | Mutated | Codeleted |
| EGD-0669 | Wild-Type | - |
| EGD-0671 | Wild-Type | - |
| EGD-0672 | Wild-Type | - |
| EGD-0675 | Wild-Type | - |
| EGD-0676 | Wild-Type | - |
| EGD-0677 | Wild-Type | - |
| EGD-0687 | Wild-Type | - |
| EGD-0688 | Mutated | Intact |
| EGD-0689 | Mutated | Codeleted |
| EGD-0691 | Wild-Type | - |
| EGD-0692 | Wild-Type | - |
| EGD-0693 | Wild-Type | - |
| EGD-0694 | Wild-Type | - |
| EGD-0700 | Mutated | Codeleted |
| EGD-0702 | Wild-Type | - |
| EGD-0703 | Wild-Type | - |
| EGD-0705 | Mutated | Intact |
| EGD-0706 | Wild-Type | - |
| EGD-0709 | Wild-Type | - |
| EGD-0710 | Wild-Type | - |
| EGD-0715 | Wild-Type | - |
| EGD-0720 | Mutated | Intact |
| EGD-0723 | Wild-Type | - |
| EGD-0725 | Wild-Type | - |
| EGD-0728 | Wild-Type | - |
| EGD-0730 | Mutated | Codeleted |
| EGD-0734 | Wild-Type | - |
| EGD-0736 | Wild-Type | - |
| EGD-0737 | Mutated | Intact |
| EGD-0738 | Wild-Type | - |
| EGD-0739 | Mutated | Codeleted |
| EGD-0742 | Wild-Type | - |
| EGD-0744 | Mutated | Intact |
| EGD-0745 | Wild-Type | - |
| EGD-0747 | Wild-Type | - |
| EGD-0753 | Wild-Type | - |
| EGD-0754 | Wild-Type | - |
| EGD-0757 | Mutated | Intact |
| EGD-0758 | Mutated | Intact |
| EGD-0759 | Wild-Type | - |
| EGD-0762 | Wild-Type | - |
| EGD-0763 | Wild-Type | - |
| EGD-0764 | Wild-Type | - |
| EGD-0765 | Wild-Type | - |
| EGD-0768 | Wild-Type | - |
| EGD-0771 | Mutated | Intact |
| EGD-0772 | Wild-Type | - |
| EGD-0773 | Mutated | Intact |
